# Ultra-rapid detection of SARS-CoV-2 in public workspace environments

**DOI:** 10.1101/2020.09.29.20204131

**Authors:** Ozlem Yaren, Jacquelyn McCarter, Nikhil Phadke, Kevin M. Bradley, Benjamin Overton, Zunyi Yang, Shatakshi Ranade, Kunal Patil, Rishikesh Bangale, Steven A. Benner

## Abstract

Managing the pandemic caused by SARS-CoV-2 requires new capabilities in testing, including the possibility of identifying, in minutes, infected individuals as they enter spaces where they must congregate in a functioning society, including workspaces, schools, points of entry, and commercial business establishments. Here, the only useful tests (a) require no sample transport, (b) require minimal sample manipulation, (c) can be performed by unlicensed individuals, (d) return results on the spot in much less than one hour, and (e) cost no more than a few dollars. The sensitivity need not be as high as normally required by the FDA for screening asymptomatic carriers (as few as 10 virions per sample), as these viral loads are almost certainly not high enough for an individual to present a risk for forward infection. This allows tests specifically useful for this pandemic to trade-off unneeded sensitivity for necessary speed, simplicity, and frugality. In some studies, it was shown that viral load that creates forward-infection risk may exceed 10^5^ virions per milliliter, easily within the sensitivity of an RNA amplification architecture, but unattainable by antibody-based architectures that simply target viral antigens. Here, we describe such a test based on a displaceable probe loop amplification architecture.

## Introduction

Once again, the world is facing a coronavirus pandemic, this time from SARS-CoV-2 that emerged in Wuhan, China in late 2019 [1]. Previous coronaviral threats include the SARS coronavirus that emerged in southern China in 2003 as the causative pathogen of severe acute respiratory syndrome [2, 3], and a coronavirus that causes Middle East respiratory syndrome (MERS) [4, 5]. However, unlike these homologous coronaviruses, SARS-CoV-2 creates a remarkable range of medical outcomes, from lethality to mild (or no) symptoms in infected individuals [6, 7]. Further, it appears to be transmissible via asymptomatic carriers. This, in turn, has caused economic disruption across the globe that is measured in the trillions of dollars [8].

At present, no single event seems likely to resolve this pandemic cleanly. Coronaviruses have generally been poor targets for vaccines [9, 10], although novel routes may make this view obsolete [11]. Antivirals that are effective against other viruses [12] may have activity against SARS-CoV-2 [13], but their ability to manage the pandemic remain in doubt. Treatments that mitigate disease symptoms may save lives, but are not likely to be effective at preventing virus spread [14, 15].

All of these factors create an urgency for tests that identify, in minutes, low- or asymptomatic infected individuals as they enter public spaces, such as workspaces, schools, points of entry, and commercial business establishments. To be useful, such tests must (a) require no sample transport, (b) require minimal sample manipulation, (c) can be performed by unlicensed individuals, (d) return results on the spot in much less than one hour, and (e) cost no more than a few dollars. They cannot involve RNA extraction or other sample preparation steps found in assays typically used in reference laboratories [16–18].

Such specs are demanding. However, the demands are mitigated by the fact that to meet its societal purpose, the test need not be ultra-sensitive. To identify carriers who have the potential to infect individuals in public spaces, tests need only be sensitive enough to catch perhaps 10^4^-10^5^ virions per sample from the nasal or oral cavities. While these numbers remain to be better defined (and on-site tests are likely to help to define them), it is clear that the necessary speed, simplicity, and frugality can be more easily obtained by trading off unneeded sensitivity. False positives are managed by re-testing. While false negatives remain (either by failure of low resource sampling or failure of the test itself), the absence of a work-place test effectively makes all untested low- or asymptomatic carries “false negatives”.

Using various nucleic acid based architectural innovations, we have developed a variety of kits that meet these specs with untrained and unlicensed users outside of traditional laboratory settings, including in the field. These have focused on detecting environmental pathogens, including detecting arboviral RNA in infected insects and ticks [19] since these require no regulatory supervision. We have established many collaborations, including TrakItNow (which builds mosquito traps) [20], immediate care facilities [21], LynxDx in Ann Arbor, MI, and Achira Labs in Bangalore, India [22].

Here, we report the application of these architectures to the detection of the SARS-CoV-2 coronavirus in nasal and oral cavity samples. The architectures that we report here:

a. Operate on dry swabbed samples, without extensive sample preparation,
b. Require no temperature cycling, and do not require expensive instruments,
c. Have ∼$3.00 in disposable costs, and therefore are routinely usable,
d. Produce an easily read signal in less than 30 minutes,
e. Have limits of detection of ∼ 200 viral RNA per assay when minimal sample preparation was sought, and
f. Are easy to run, not requiring a trained medical professional.

## Material and Methods

### 1. Primers and displaceable probes for displaceable probe RT-LAMP (DP-RT-LAMP)

Primer design was performed using in-house software (OligArch v2) to create primer sets that account for the evolutionary variation within the genomes of the target viruses. Viral sequences for SARS-CoV-2 and other beta-coronaviruses that infect humans (SARS, MERS, HKU1, and OC43) were downloaded from the NIAID Virus Pathogen Database and Analysis Resource (ViPR) [23]. Multiple sequence alignments (MSAs) were created for these sequences using MUSCLE v3.8.31 [24]. The resulting MSAs were used as input to OligArch v2, which searches for primer sets that are conserved within a target of interest while avoiding unintended targets also included within the MSA (here allowing distinction between SARS-CoV-2 and other coronavirus types).

Rules for DP-RT-LAMP design were adapted using criteria from the Eiken Genome website [25]. Designed DP-RT-LAMP sets were compared to the NCBI RNA virus database using NCBI BLAST [26] to eliminate sets that might cross-react. Sets were further compared, using in-house software PrimerCompare v1, to eliminate sets with primers that would dimerize in a multiplexed assay to produce the final sets of DP-RT-LAMP primers.

The DP-RT-LAMP primers and strand displaceable probes were purchased from Integrated DNA Technologies (IDT, Coralville, IA). Strand-displaceable probes were 5’-quencher labeled with Iowa Black-FQ (IBFQ). Fluorescently labeled displaceable probes partially complementary to the quencher labeled probes were 3’-labeled with FAM. Alternatively, for multiplexing purposes, internal control probes targeting the human RNase P gene were 5’-labeled with IBFQ and 3’-labeled with JOE in addition to existing probe with FAM-label. The double strand portion of the probes were screened against any viral genome and human genomic sequence.

### 2. SARS-CoV-2 templates

#### IVT RNA fragment preparation

Target RNA was generated from synthetic DNA fragments of the viral genes of interest. Synthetic DNA gene fragments were ordered from IDT as gBlocks. An initial PCR introduced the T7 promoter. Next, 150 nM of PCR product was used in T7 RNA transcription reaction (50 µL total volume); the reaction mixture was incubated at 37°C for 16h. DNA templates were removed by digestion with DNase I, the mixture was phenol-CHCl3 extracted, and the RNA was recovered by EtOH precipitation. The product RNA was quantified using a Nanodrop UV spectroscopy, and reference materials with known concentrations were prepared in serial dilutions in TE buffer (10 mM Tris pH 7.0, 1 mM EDTA) and aliquots were stored at −80°C.

#### Fully synthetic SARS-CoV-2 RNA

Synthetic SARS-CoV-2 RNA Control was from Twist Bioscience (MT007544.1, 1×10^6^ RNA copies/µL). It was used for initial limit of detection (LOD) studies. Appropriate dilutions were made in 1 mM Na citrate pH 6.5, 0.4 U/µL RNase inhibitor (NEB, Ipswich, MA) and aliquots were stored at −80°C.

#### Heat-inactivated SARS-CoV-2 isolate

Authentic SARS-CoV-2, isolate USA-WA 1/2020, was obtained through BEI Resources (cat no. NR-52286, 1.16×10^9^ genome equivalents/mL). This virus has been inactivated by heating at 65°C for 30 minutes. Target dilutions were made in 1 mM Na citrate pH 6.5, 0.4 U/µL RNase inhibitor (NEB, Ipswich, MA) and aliquots (100 µL) were stored at −80°C. This target was used to determine final LODs and spike-in experiments where minimal sample preparation methods were sought for nasal swab and saliva sampling.

### 3. DP-RT-LAMP Assay

12.5 µL of 2X WarmStart LAMP master mix (NEB) was combined with 2.5 µL of 10X LAMP primer set, 1 µL of excess B3 primer (300 µM), 0.2 µL of dUTP (100 mM, Promega), 0.5 µL of Antarctic Thermolabile UDG (1U/µL, NEB), 0.5 µL of RNase inhibitor (40U/µL, NEB), 2 µL of template RNA or inactivated virus isolate, and 6 µL of nuclease-free water (or briefly processed nasal/saliva samples) to bring the final reaction volume to 25 µL.

10X LAMP primer set consists of 16 μM each of FIP and BIP, 2 μM each of F3 and B3, 5 μM LF (or LB for CoV2-v2-4 set), 4 µM LB (or LF for CoV2-v2-4 set), 150 nM quencher-bearing probe, and 100 nM of fluorophore-bearing probe.

Reactions were monitored in real-time using either a LightCycler^®^ 480 (Roche Life Science, US) or a Genie^®^ II (Optigene, UK) instrument. 8-strip PCR tubes were first incubated at 55°C for 10 min followed by incubation at 65°C for 45-60 min. During the 65°C incubation, fluorescence signal was recorded every 30 seconds using FAM/SYBR channel of the instrument. End-point observation of the fluorescence signal generated by strand displaceable probes was enabled by blue LED light (excitation at 470 nm) through orange filter of SafeBlue Illuminator/ Electrophoresis System, MBE-150-PLUS (Major Science, US) or 3D printed observation box (Firebird Biomolecular Sciences, US).

## 4. From brief sample preparation to DP-RT-LAMP testing

### Ethical Statement

Nasal swabs and saliva samples were collected from healthy volunteers, as approved by IntegReview IRB procedure (protocol number: 2020001). Collected samples were then spiked in with heat-inactivated SARS-CoV-2 virus at varying concentrations. Informed consent was obtained for all participant samples.

### 4.1 Mid-turbinate and anterior nasal swab sampling

#### Sample collection

CleanWIPE Swab, 3” Semi-Flexible bulb tip (HT1802-500, Foamtec International) was used for nasal sampling. Each nostril was swabbed for at least 10 seconds using the same swab. Swab was placed in sterile 15 mL falcon tube and stored at 4°C until processing. Swabs were processed within 1 hour.

#### Sample preparation

Nasal swab was eluted in 200 µL of buffer solution (1 mM Na citrate pH 6.5, 2.5 mM TCEP, 1 mM EDTA, 10 mM LiCl, 15% Chelex-100) by brief vortexing. Swabs were then removed and elution solution was briefly spun down. 6 µL of sample elution was combined with 2 µL of inactivated virus (BEI resources) or water (no template control) and added to 17 µL of DP-RT-LAMP reaction mixture (12.5 µL of 2X WarmStart LAMP master mix, 2.5 µL of 10X LAMP primer set, 1 µL of excess B3 primer (300 µM), 0.2 µL of dUTP (100 mM,), 0.5 µL of Antarctic Thermolabile UDG (1U/µL, NEB), 0.5 µL of RNase inhibitor (40U/µL, NEB). Samples were then incubated and analyzed in real-time as described above. A DP-RT-LAMP assay for detecting RNase P was run in parallel as a control to ensure the sufficient sample collection.

#### Other sample preparation methods tested

We initially tested TE buffer (10 mM Tris-HCl pH 7.0, 1 mM EDTA) for eluting nasal swabs. Another method involved use of an inactivation buffer containing 10 mM NaOH, 2.5 mM TCEP and 1 mM EDTA followed by incubation at 95°C for 5 min [27]. Same buffer solution was then coupled with 15% Chelex-100 and extended heating (56°C 15 min and 95°C 5 min).

### 4.2 Saliva as a sample for DP-RT-LAMP

Saliva samples were either collected before brushing the teeth or 1h after brushing the teeth. 1 mL of saliva was collected in sterile 5 mL falcon tube and stored at 4°C until processing and samples were processed within 1h.

In addition to IRB approved saliva collection, some of the saliva samples were purchased from BioIVT (human saliva, 5 mL, gender unspecified.)

#### Sampling procedure

A suspension (100 µL) of 15% Chelex-100 in a 1.6 mL microcentrifuge tube was spun down briefly and supernatant was removed. To this was added 100 µL of saliva mixed with 1 µL of concentrated sample preparation solution (0.1 M Na citrate pH 6.5, 1M LiCl, 0.25 M TCEP, 0.1 M EDTA). Each sample was briefly vortexed and spun down to settle the Chelex-100. The saliva sample (6 µL) was combined with 2 µL of inactivated virus (BEI resources) or water (no template control) and added to 17 µL of displaceable probe RT-LAMP mixture. The reaction was allowed to proceed as previously described. A LAMP assay for detecting RNase P gene was run in parallel as a control to ensure the sufficient sample collection.

### 4.3 Collecting saliva on Q-paper and DP-RT-LAMP testing

#### Q-paper preparation

Whatman filter paper (1g) was immersed in 50 mL of 1.8% aq. NaOH solution for 10 min. Treated paper was collected by filtration and immersed in aq. EPTMAC (2,3-epoxypropyl) trimethylammonium chloride) solution for 24h at RT. The mass ratio of EPTMAC to filter paper was 0.28 to 1. Cationic (Q) paper was collected by vacuum filtration and neutralized with 50 mL of 1% AcOH. Final product was washed three times with ethanol (96%) and dried at 55°C for 1h. Q-paper sheets were cut into small rectangles (∼ 0.5 x 0.2 cm) for saliva collection.

#### Saliva sampling

Q-paper was first dipped into saliva samples and soaked for 5 seconds, then air dried for 5 min. Q-paper containing saliva was directly inserted into 50 µL of DP-RT-LAMP mixture (25 µL of 2X WarmStart LAMP master mix, 5 µL of 10X LAMP primer set, 2 µL of excess B3 primer (300 µM), 0.35 µL of dUTP (100 mM,), 1 µL of Antarctic Thermolabile UDG (1U/µL, NEB), 1 µL of RNase inhibitor (40U/µL, NEB) and 16 µL of nuclease-free water) and reaction was allowed to proceed as described above. A LAMP assay for detecting RNase P gene was run in parallel as a control to ensure the sufficient sample collection.

## 5. Clinical nasopharyngeal (NP) samples

### Ethical Statement

Samples were collected and tested under a protocol reviewed and approved by GenePath Dx / Causeway Healthcare’s Independent Ethics Committee/ Institutional Review Board which is registered with the Central Drugs Standard Control Organization (CDSCO), Office of Drugs Controller General (DCG), Directorate General of Health Services, Ministry of Health and Family Welfare (MoHFW), Government of India (with registration number ECR/225/Indt/MH/2015). Informed consent was obtained for all participant samples.

### Sample preparation

NP swabs, previously stored in VTM media, were eluted in 200 µL of buffer solution (1 mM Na citrate pH 6.5, 2.5 mM TCEP, 1 mM EDTA, 10 mM LiCl, 15% Chelex-100) by brief vortexing. Swabs were then removed and elution solution was briefly spun down. 5 µL of sample was combined with 20 µL of RT-LAMP reaction mixture (12.5 µL of 2X WarmStart LAMP master mix, 2.5 µL of 10X LAMP primer set, 1 µL of excess B3 primer (300 µM), 0.2 µL of dUTP (100 mM,), 0.5 µL of Antarctic Thermolabile UDG (1U/µL, NEB), 0.5 µL of RNase inhibitor (40U/µL, NEB and 3 µL nuclease-free water). A LAMP assay for detecting RNase P gene was run in parallel as a control to ensure the sufficient sample collection.

Reactions were monitored in real-time using Rotor Gene Q (Qiagen, US). Samples were first incubated at 55°C for 10 min followed by incubation at 65°C for 45 min. During the 65°C incubation, fluorescence signal was recorded every 30 seconds using FAM channel for SARS-CoV-2 detection and JOE channel for RNase P detection.

## 6. Multiplexed DP-RT-LAMP to detect SARS-CoV-2 and RNase P (Internal Control)

### BEI template in human RNA background

Varying amounts (10^5^, 10^4^, 10^3^, 10^2^, 10 and 5 copies) of heat-inactivated SARS-CoV-2 (BEI resources) spiked with 440 copies of human RNA, 1.25 µL of 10X CoV2-W3 LAMP primer set (FAM-labeled probe) and 1.25 µL of 10X RNaseP-2 LAMP primer set (JOE-labeled probe) were added to RT-LAMP mixture (25 µl total volume).

Multiplexed reaction mixtures were pre-incubated at 55°C for 10 min, then 65°C for 50 min and the fluorescence signals from three channels were recorded every 30 seconds using LightCycler^®^ 480 (Roche Life Science, US) during 65°C incubation. Channel 483-533 is specific to FAM-labeled SARS-CoV-2 probe, channel 523-568 is an intermediate channel detecting signals from both FAM- and JOE-probes, and channel 558-610 is specific to JOE-labeled RNase P probe.

### BEI template spiked into nasal swab or saliva samples

A briefly processed nasal swab or saliva sample (∼6 µL) from Section 4.1 and 4.2, respectively, was combined with 2 µL of heat-inactivated SARS-CoV-2 isolate to give 10^4^, 10^3^, 10^2^ or 0 RNA copies per assay. Some samples were heat treated at 95°C for 5 min prior to addition of RNA template. Samples with viral RNA were then added to RT-LAMP mixture containing both CoV2-W3 and RNAseP-2 LAMP primers in equal amounts (1.25 µL each of 10X LAMP primer sets) to give a total of 25 µL assay volume. Real-time analysis was performed as mentioned above using LightCycler^®^ 480 with three fluorescence channels.

## 7. Lyophilization of DP-RT-LAMP reagents

### Dialysis example for 10 LAMP reactions

Bst 2.0 WarmStart^®^ DNA Polymerase (10 µL, 8U/µL, NEB), 10 µL of WarmStart^®^ RTx Reverse Transcriptase (15 U/µL, NEB), 5 µL of Antarctic Thermolabile UDG (1U/µL, NEB), 5 µL of RNase inhibitor (40U/µL, NEB) was combined with 170 µL of dialysis buffer (10 mM Tris-HCl pH 7.5, 50 mM KCl, 1 mM DTT, 0.1 mM EDTA, 0.1% Triton X-100). 200 µL mixture was then placed in an ultrafiltration membrane (10 kDA cut-off limit, Millipore, Billerica, MA). Samples were centrifuged at 13,000 rpm for 8 min, then further washed with 250 µL of dialysis buffer twice to concentrate resulting glycerol free enzyme mix down to 30 µL. 30 µL of enzyme mix was combined with 25 µL of 10X LAMP primer set, 10 µL of 300 µM B3 primer, 35 µL of dNTP mix (10 mM each of dATP, dCTP, dGTP and 5 mM each of dTTP and dUTP) and 25 µL of 1M D-(+)-trehalose. Combined mixture was then distributed into 8-strip PCR tubes as 12.5 µL aliquots. Samples were frozen by liquid nitrogen and lyophilized for 4-6h. Lyophilized reagents were stored at RT and tested within 7 days.

### Reconstitution of lyophilized DP-RT-LAMP reagents

Sample (6 µL, nasal swab/saliva or RNA template) was mixed 19 µL of reconstitution buffer (2.5 µL 10X isothermal amplification buffer (NEB), 1.5 µL 100 mM MgSO4 and 15 µL of nuclease-free water) and added into lyophilized reagents. RT-LAMP reactions were monitored in real-time using Genie II and fluorescence signal was visualized as described in previous sections.

## Results

### Assay architecture

In recent years, reverse transcription loop-mediated isothermal amplification (RT-LAMP) has become an alternative to RT-PCR due to its high sensitivity and specificity, its tolerance for inhibitory substances, and operation at constant temperatures. Together, these lower assay complexity and cost, making LAMP often considered for COVID-19 diagnostics [28–30].

In its classical form, RT-LAMP uses six primers binding eight distinct regions within a target RNA (**Fig 1**). It runs at constant temperatures ranging from 62 °C to 72 °C, and uses a reverse transcriptase and a DNA polymerase with strong strand displacing activity (e.g. Bst DNA polymerase). During the initial stages of RT-LAMP, forward and backward internal primers (FIP and BIP), with outer forward and backward primers (F3 and B3), form a double loop structure. This becomes the seed structure for subsequent LAMP amplification. Amplification rate is further improved by two loop primers (LF and LB), which are designed to bind the single stranded regions of the loops. These yield concatemers with multiple repeating loops [31].

**Fig 1.**
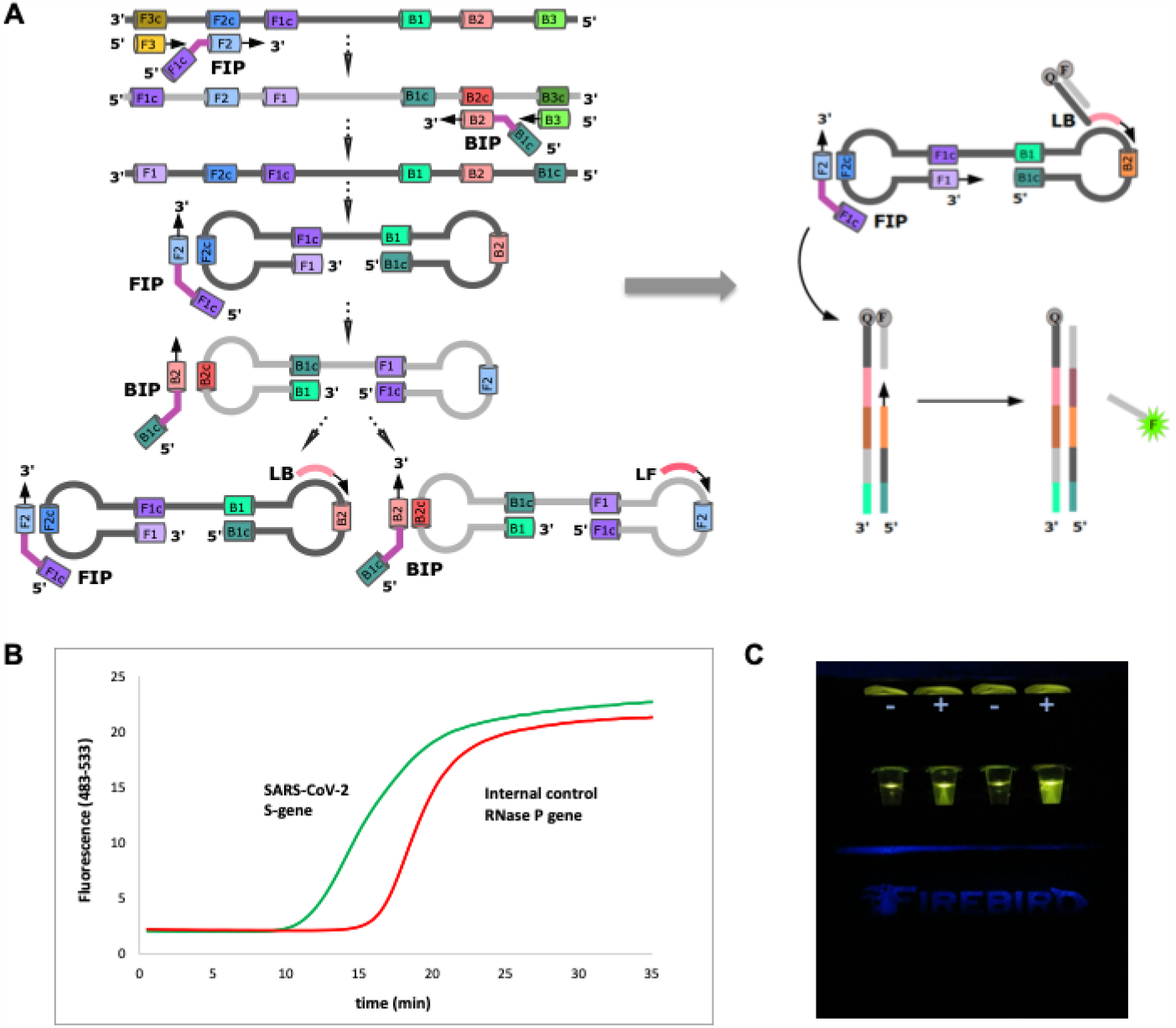
Displaceable probe LAMP to detect SARS-CoV-2. **(A)** RT-LAMP is initiated by adding internal primers (FIP or BIP) that annealed to F2c or B2c regions. Outer primer (F3 or B3) then hybridizes to F3c or B3c and initiates the formation of self-hybridizing loop structures by strand invasion of the DNA sequences already extended from the internal primers (FIP and BIP). The resulting dumbbell structure is then used as a seed for exponential LAMP amplification by a strand displacing polymerase. This process is further accelerated by the introduction of loop primers (LF or LB) hybridizing to segments between F1c and F2 or B1c and B2, respectively. Further, priming region of the quencher labeled probe (e.g. LB) is extended by a strand-displacing polymerase, and primer extension from the reverse primers then reads through the primer on the quencher labeled probe, displacing the probe that bears the fluorophore. This architecture differs from standard LAMP in how the signaling element moves in the detection architecture **(B)** This results in increase in the fluorescent signal and real-time analysis of the process manifests itself as sigmoidal curve as it would be in RT-qPCR using TaqMan probes. **(C)** In addition to real-time analysis, end-point fluorescence can be visualized using an observation box with blue LED exciting at 470 nm through orange filter (Firebird Biomolecular Sciences LLC, US).

Classical LAMP generates signals by the precipitation of the magnesium salt as one of its byproducts, pyrophosphate; the turbidity from this precipitation is detected. Alternatively formation of high molecular weight amplicons allows an intercalating dye to create a fluorescent signal [32, 33] or non-fluorescent signal [34, 35]. Alternatively, the pH change arising during the amplification is detected by the change in the color of an indicator [36–38].

None of these are well suited for workplace detection of pathogen RNA, such as that from SARS-CoV-2. These detection architectures can easily be deceived by off-target amplicons, and are therefore susceptible to generation of false-positive results. Confirming the nature of the amplicon by measuring melting temperatures, very useful in PCR, is difficult with LAMP amplicons whose lengths mature over the time of the process, and where off-target amplicons have unpredictable melting temperatures [39–41]. Assimilating probes have been introduced to allow the high molecular weight amplicon to contain a fluorophore, where the assimilation separates the fluorophore from a fluorescence quencher [42].

To manage these issues, we offered an alternative architecture that exploits a displaceable probe (DP). This is a short oligonucleotide carrying a 3’-fluorophore that is displaced from a complementary oligonucleotide as the desired amplification is completed. That complementary oligonucleotide has a 5’-quencher, and carries a tag that is a primer that binds to one of the loops in the initial LAMP double loop structure (**Fig 1A**). Thus, each probe is delivered to the amplification mixture as a target-sequence-independent double-strand probe region and a single-stranded target-priming region. The sequences of all components are shown in **Table 1**.

**Table 1.**
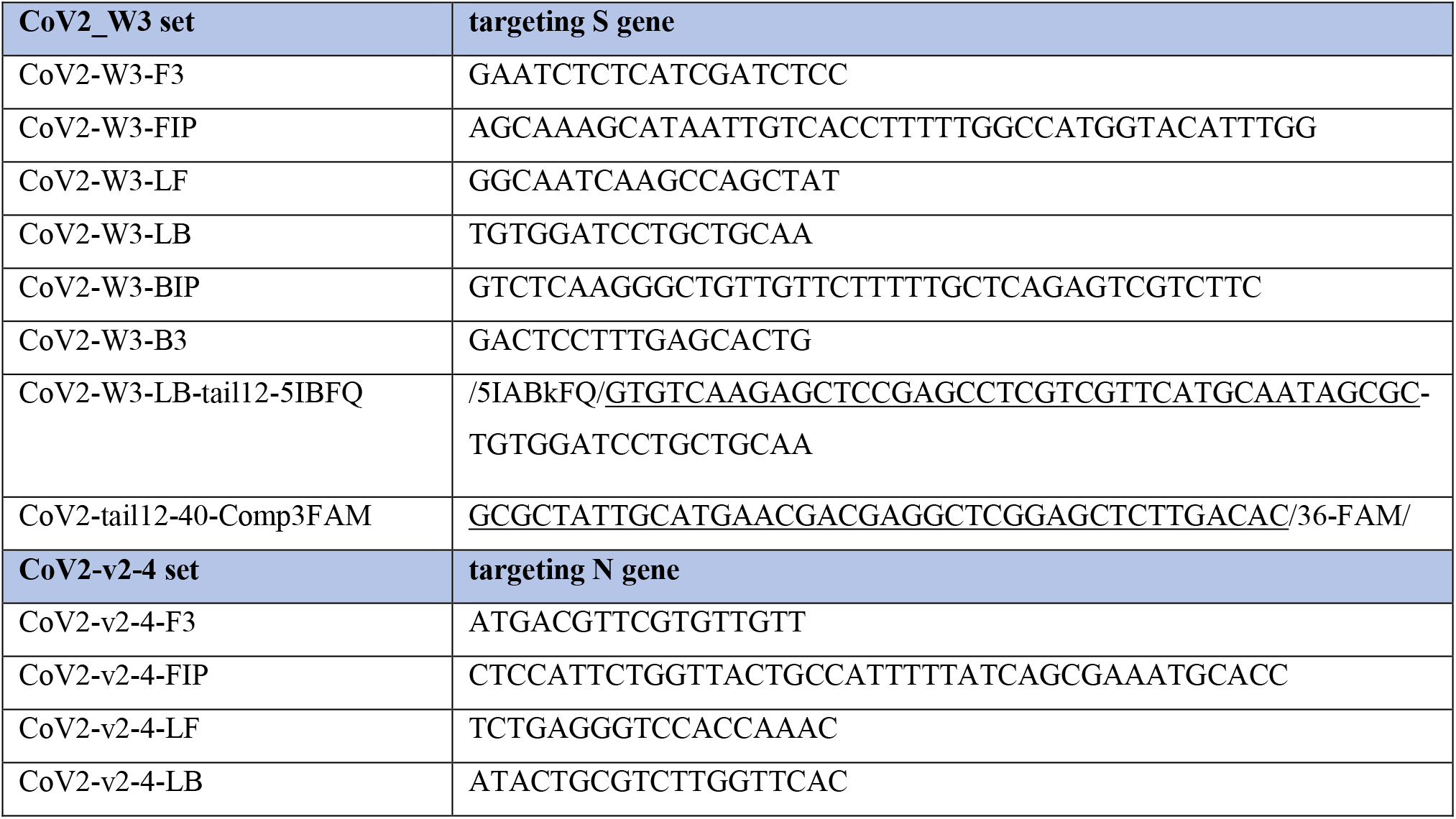
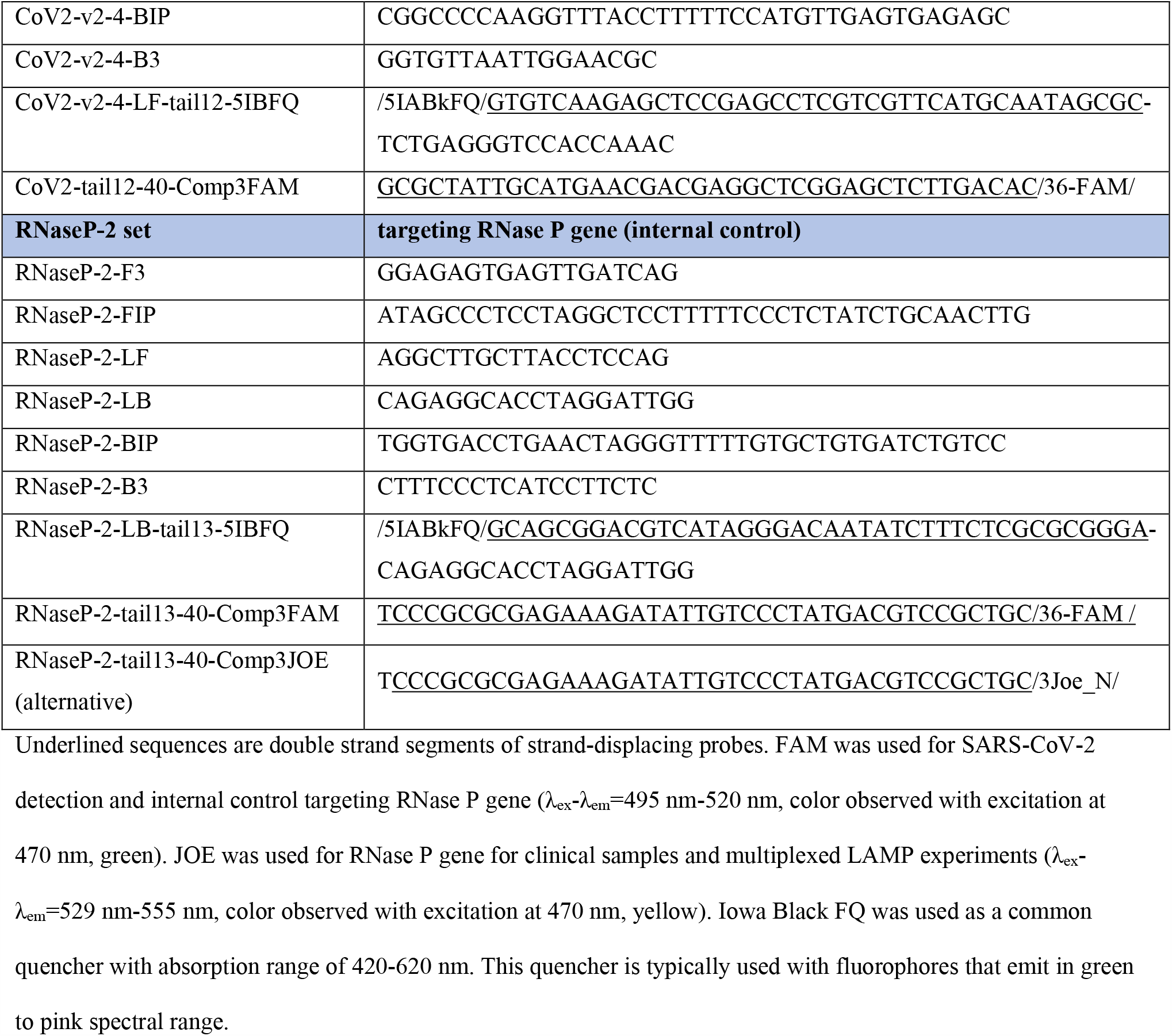
LAMP primers and probe sequences used in this study.

In the displaceable probe architecture, in the absence of target, no fluorescence is observed due to quenching of fluorophore by a quencher in the un-displaced duplex. In the presence of target, the single-stranded portion of the quencher probe binds to the target and thus extended. Further polymerase extension by reverse primers displaces the quencher strand from the fluorescently labeled strand, allowing the emission of fluorescence and its analysis in real-time. As a consequence of the displacing process, “S-shaped” curves appear in a plot of fluorescence versus time, similar to RT-PCR and similar Ct (or Tt-threshold time) analyses **(Fig 1B)**.

With this architecture, displaceable probes are fluorophore labeled whose sequences are unrelated to the sequence of the target analyte, and to be released only after the amplification fully starts. This allows totally independent selection of the duplex sequence. This, in turn allows it to be captured, either downstream or while the amplification is occurring. Signal arising from the displaced probes are typically generated within 20 min and visible to human eye. Signals can be visualized with an observation box that uses a blue LED and an orange filter **(Fig 1C)**. This box is fabricated by 3-dimensional printing.

### Sensitivity of DP-RT-LAMP assay using IVT RNA and Twist RNA

Our first experiments sought to measure the sensitivity of a specific RT-LAMP primer set (CoV2-W3) that had been selected from three trial sets that targeted the spike region of the virus genome. Here, RNA targets were prepared by transcription of a DNA template (230 nt). Varying concentrations of RNA were used to determine assay sensitivity; assay conditions (65°C, 60 min) followed those established to detect a panel of RNA viruses in mosquitoes [17].

With this target, limits of detection (LODs) were 5 copies/assay, giving a threshold time (Tt, equivalent of Ct) of 22.5 min **(S1 Fig)**. When the synthetic RNA target was replaced by the complete RNA genome (Twist Biosciences, SARS-CoV-2 RNA), the sensitivity dropped to 100 copies/assay with Tt = 25.3 min **(S2 Fig)**.

We then sought conditions to increase the sensitivity of the assay. These included:

a. adding a second reverse transcriptase (SuperScript IV (SSIV) to the WarmStart reverse transcriptase (WS-RTx, NEB) already present.
b. changing the reaction buffer,
c. adding random hexamers (12 µM)
d. adding excess reverse primer (B3 primer), and
e. varying the incubation temperature (**S1 Table and S2 Table**).

Each reaction mixture was pre-incubated at 55°C for 10 min to ensure formation of sufficient cDNA by the reverse transcriptase. This was then followed by incubation at 65°C.

These modifications improved sensitivity with the full-length RNA genome; LODs improved to 10 copies/assay. This compares favorably with SARS-CoV-2 colorimetric assay from New England Biolabs, which has a reported LOD of 500 copies/assay [36]. However, use of 5X SSIV buffer resulted in fluorescent signal in the absence of target (no template controls, NTCs). This drove the choice of the presently preferred conditions that (i) use the original NEB buffer, (ii) WS-RTx as the only reverse transcriptase, (iii) in the presence of random hexamers, and (iv) or excess B3 primer. These conditions gave no “NTC problem” up to 60 minutes, with an LOD of 10 copies/assay. Refining these conditions further, better Tt values were observed with excess B3 than with random hexamers. Therefore, excess B3 primer was used in further RT-LAMP experiments.

### DP-RT-LAMP assays with high sensitivity using heat-inactivated virus isolate

We then assessed the LOD with authentic, non-synthetic virus that had been heat-inactivated (SARS-CoV-2 isolate, inactivated at 65°C for 30 minutes, BEI Resources). This was also used to “spike” nasal swabs and saliva samples. Three conditions previously tested for the synthetic RNA (Twist Bioscience) were again tested for the BEI target. The best sensitivity (10 copies/assay) was achieved with “Condition 1”, using the NEB isothermal amplification buffer, excess B3 primer, and WS-RTx with incubation at 55°C for 10 min (initially), followed by further incubation at 65°C 50 min (**S3 Fig**).

With the current modifications in the RT-LAMP protocol, another DP-RT-LAMP primer set was designed to target the N gene of SARS-CoV-2. We also designed a DP-RT-LAMP primer set to targeted the human RNase P gene. Detection of the amplicon from human RNase P was intended to serve as an internal control to assess the adequacy of the sample collection.

The primer set targeting S gene (CoV2-W3) gave an LOD of 10 copies/assay within 16 min (**Fig 2A**); the fluorescence signal arising from fluorescence was excited at 470 nm (typically an LED) and visualized through an orange filter to block the excitation light (**Fig 2B**). The primer set targeting the N gene (within the BEI sample) had an LOD of 25 copies/assay within a 12 min (**Fig 2C**). The system targeting the human RNase P gene had an LOD of 44 copies/assay, within a 16 min (**Fig 2D**). Threshold times were compared to RT-qPCR where N gene and RNase P gene were detected in multiplex format (Yang et al., *manuscript in preparation*). For this comparison, Ct values from PCR assay were converted to their corresponding Tt values; RT-LAMP was found to be outperforming over multiplex RT-qPCR in terms of assay rapidity (**Fig 2E**).

**Fig 2.**
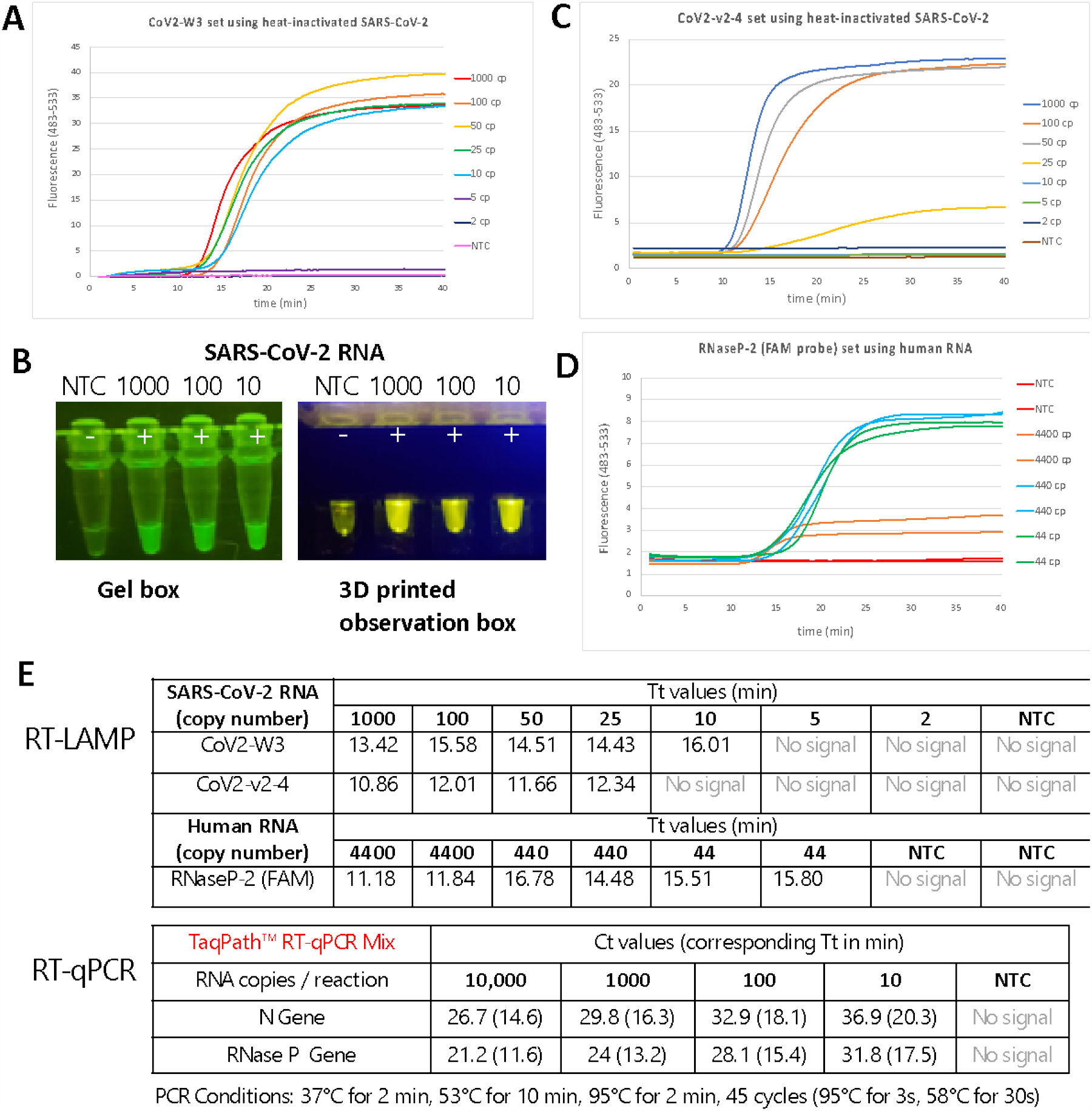
Limit of detection using DP-RT-LAMP primers using heat-inactivated SARS-COV-2 or human RNA (for internal control). **(A)** Real-time analysis of CoV2-W3 primer set (targeting S gene) showed that LOD was 10 copies of RNA/assay. **(B)** End-point visualization of LAMP products with primer set CoV2-W3 using SafeBlue Illuminator/Electrophoresis System or using a hand-held observation box with integrated blue LED and orange filter (Firebird Biomolecular Sciences LLC). **(C)** Real-time analysis of CoV2-v2-4 primer set (targeting N gene) showed an LOD of 25 copies of RNA/assay. **(D)** Real-time analysis of internal control RNaseP-2 primer set (targeting human RNase P gene) showed that LOD was 44 copies of human RNA/assay. **(E)** Time to threshold (Tt) values of each LAMP primer set was determined for each target copy number/assay and similar values were obtained when compared to RT-qPCR test (Ct values were also converted to their corresponding Tt values for convenience).

### Simple sample preparation of nasal swabs and saliva samples

For a test that can be used at the entrance to a public space to identify carriers who present an environmental risk, sample preparation must be minimal, and any instrumentation involved must be “field-deployable”. To be fool-proof, end-point analysis is demanded. Several research groups have also sought low sample preparation workflows, as RNA purification from biological samples is time consuming and timely delivery of test results can be impaired due to limited supplies of sample purification kits [27–29, 43–45]. To meet these specs, we generated three protocols for SARS-CoV-2 testing.

First, the behavior of the virus itself defines the sampling procedure. False negatives arising from defective sampling are often as problematic as (or more problematic than) false negatives arising from failure of the assay. Fortunately, the life cycle of SARS-CoV-2 appears to allow simple sampling, with mid-turbinate sampling being adequate, as well as saliva sampling [46, 47].

Therefore, our first protocol uses dry mid-turbinate or anterior nasal swabbing as a collection method, and relied on the positive control targeting human RNase P to ensure that the collection was adequately aggressive. Post sampling, swabs were eluted in various elution/ inactivation buffers. An aliquot from the elution solution was added directly to the DP-RT-LAMP mixture, and analyzed in real-time and by visualization of end-point fluorescence (**Fig 3A**). Spiked saliva (with saliva alone as the negative control) diluted with concentrated inactivation buffer (1:100 ratio of buffer to saliva) was also used. An aliquot of the resulting mixture was added to the DP-RT-LAMP mixture and analyzed similarly (**Fig 3B**).

**Fig 3.**
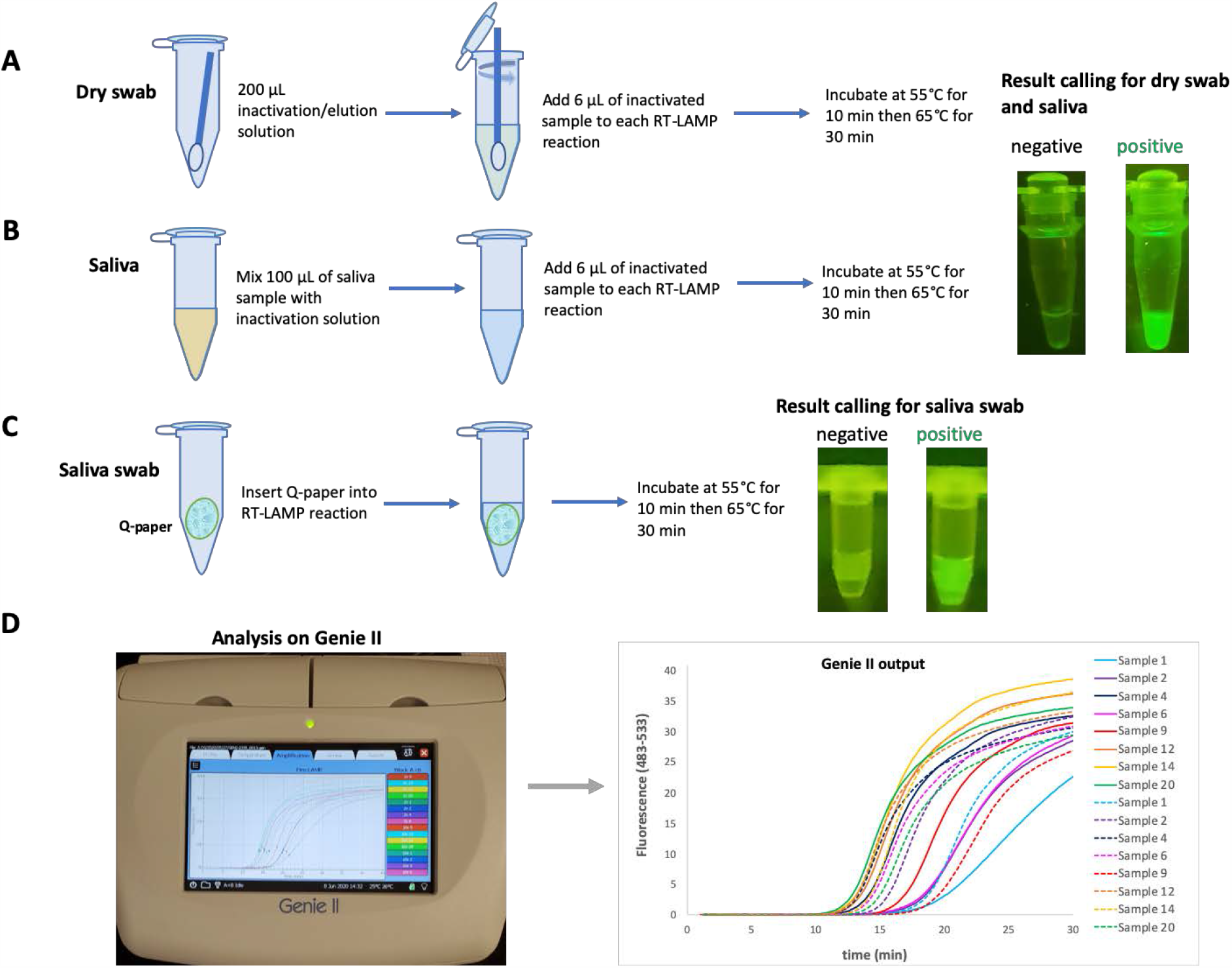
Sampling work-flow and results output. **(A)** Dry nasal swabs were used as sampling method. Swabs were first eluted in a sample preparation buffer and aliquot from that was added into RT-LAMP mixture. End-point results were visualized using blue LED and orange filter. **(B)** Direct saliva was mixed with a sample preparation buffer briefly and aliquot from that was added into RT-LAMP mixture. End-point results were visualized using the same method for nasal swab sampling. **(C)** Quaternary ammonium modified paper (Q-paper) was combined with saliva and Q-paper coated with saliva was directly introduced into RT-LAMP mixture without further manipulation. End-point fluorescent signal was visualized using blue LED and orange filter. Note that the square of Q-paper is observable, but does not compromise the real-time or end-point analysis. **(D)** In addition to end-point visualization, RT-LAMP experiments were also run in real-time using Genie^®^ II (Optigene, UK) which can operate on battery therefore enabling its use in low-resource settings.

Alternatively, saliva can be placed on “Q-paper”, a cellulose filter paper that carries quaternary ammonium groups. Q-paper has been previously used to capture arboviral RNA from single mosquitoes after a drop of ammonia is added to the carcasses [17]. In this work, the Q-paper holding the viral RNA could be added directly to the RT-LAMP mixture without any sample preparation. The fluorescence can be analyzed in real-time or by end-point visualization, again using blue LED excitation with fluorescence observed through an orange filter (**Fig 3C**). The fluorescence can also be seen in a hand-held observation box.

Real-time analyses of all methods tested were performed on a (Roche LightCycler^®^ 480), which is not easily portable. However, performance was equally satisfactory when done on a portable Genie^®^ II instrument, available from Optigene. Genie^®^ II processes 16 samples simultaneously using the FAM-channel (483-533 nm). The data outputs are similar to those obtained with the more expensive real-time PCR instrument. Genie^®^ II offers positive/negative results with Tt values as good as obtained with the PCR instrument, but at a fraction of the cost and useable in the lobby of a workplace, a courtroom, or a school (**Fig 3D**).

### Validation of DP-RT-LAMP assay with contrived nasal swabs

Having established work-flow parameters, we tested various elution/inactivation buffers with or without a heat step to design the presently preferred protocol. **Fig 4A** summarizes the methods used to process mid-turbinate or nasal anterior swabs. TE (Tris-HCl pH 7.0 and 1 mM EDTA) as an elution buffer gave LODs ≈ 1000 copies/assay, with Tt values of ≈30 min. The procedure of Rabe and Cepko [27] was used, with swabs eluted in buffer containing NaOH, TCEP and EDTA and incubated at 95°C for 5 min, and then spiked with known concentrations of BEI template. These gave LODs as low as 100 copies/assay, with a Tt of 23.5 min. Addition of 15% Chelex-100 to Rabe and Cepko method improved the Tt by 2 min.

**Fig 4.**
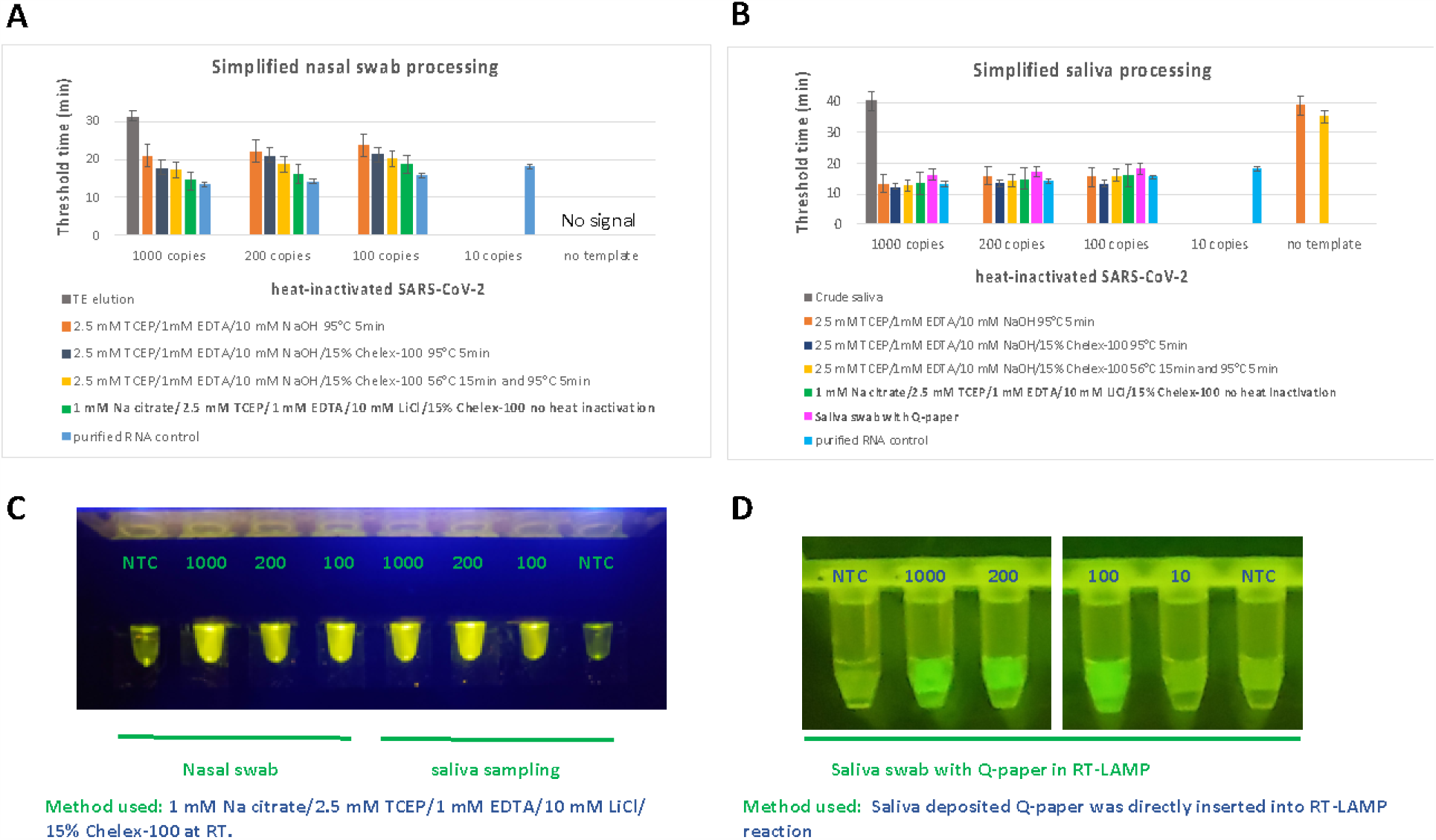
Optimization of sampling methods and fluorescence visualization with presently preferred methods. **(A)** Five different methods were evaluated for nasal swab sampling, including (i) TE elution, (ii) the method of Rabe-Cepko *et al*., (iii) a method combining Cepko with Chelex-100, (iv) a method combining Cepko with Chelex-100 with two-step heating, and (v) a process without a heating step. Heat-inactivated SARS-CoV-2 isolate was spiked into nasal swab elutions and each method’s sensitivity was determined. Purified RNA control was included as a reference. **(B)** For saliva sampling, six methods were evaluated: (i) crude saliva without any treatment, (ii) the Cepko method, (iii) Cepko method coupled with Chelex and a heat-step, (iv) Cepko method with Chelex and two-step heating, (v) a process without a heating step, and (vi) deposition of saliva on Q-paper and its direct introduction into RT-LAMP. A purified RNA control was included as a reference. **(C)** End-point visualization of finalized methods: Nasal swab and one of the saliva sampling methods uses buffer solution containing 1 mM Na citrate pH 6.5, 2.5 mM TCEP, 1 mM EDTA, 10 mM LiCl and 15% Chelex-100. LODs for both samplings were determined to be 100 copies/assay **(D)** End-point visualization of saliva deposited on Q-paper and its direct use in DP-RT-LAMP reaction. The LOD was 100 copies/assay using Q-paper.

Despite its promise, this approach did not give reproducible results when nasal swabs were spiked with inactivated virus prior to the 95°C heating step. A similar problem was observed when same buffer was combined with Chelex-100, in a workflow that incorporated two heating steps, one at 56°C for 15 min and a second at 95°C for 5 min. Dao Thi *et al*. [44] also report similar results when nasal swab elution mixtures were spiked with RNA, and then heated (95°C, 5 min). However, processing of clinical samples using the method developed by Rabe and Cepko [27] with heating at 95°C for 5 min did not cause a decrease in assay sensitivity [43].

Seeking to further simplify sampling work-flow, we modified the elution by replacing NaOH with sodium citrate (pH 6.5) and added TCEP, EDTA, LiCl and Chelex-100. Swabs were eluted at room temperature without any additional heating step. Here, 100 copies of viral RNA were detectable per assay within 18 min, only delayed by 2 min when compared to BEI control template. In addition to real-time analysis, end-point fluorescent images were also visible to human eye at 100 copies/assay (**Fig 4C**).

The sensitivity of the current nasal swab sampling method was analyzed with a larger number of samples using contrived nasal swabs of healthy volunteers. Here, 1000 RNA copies/assay were detected consistently at 100%. Ca. 200 copies/assay were detected with 90% efficiency, and 100 copies of RNA/assay were detected at 50% efficiency. The internal control that targets the RNase P gene was detected at 100%, indicating that the sample collection was sufficiently aggressive (**Fig 5A and 5D**).

**Fig 5.**
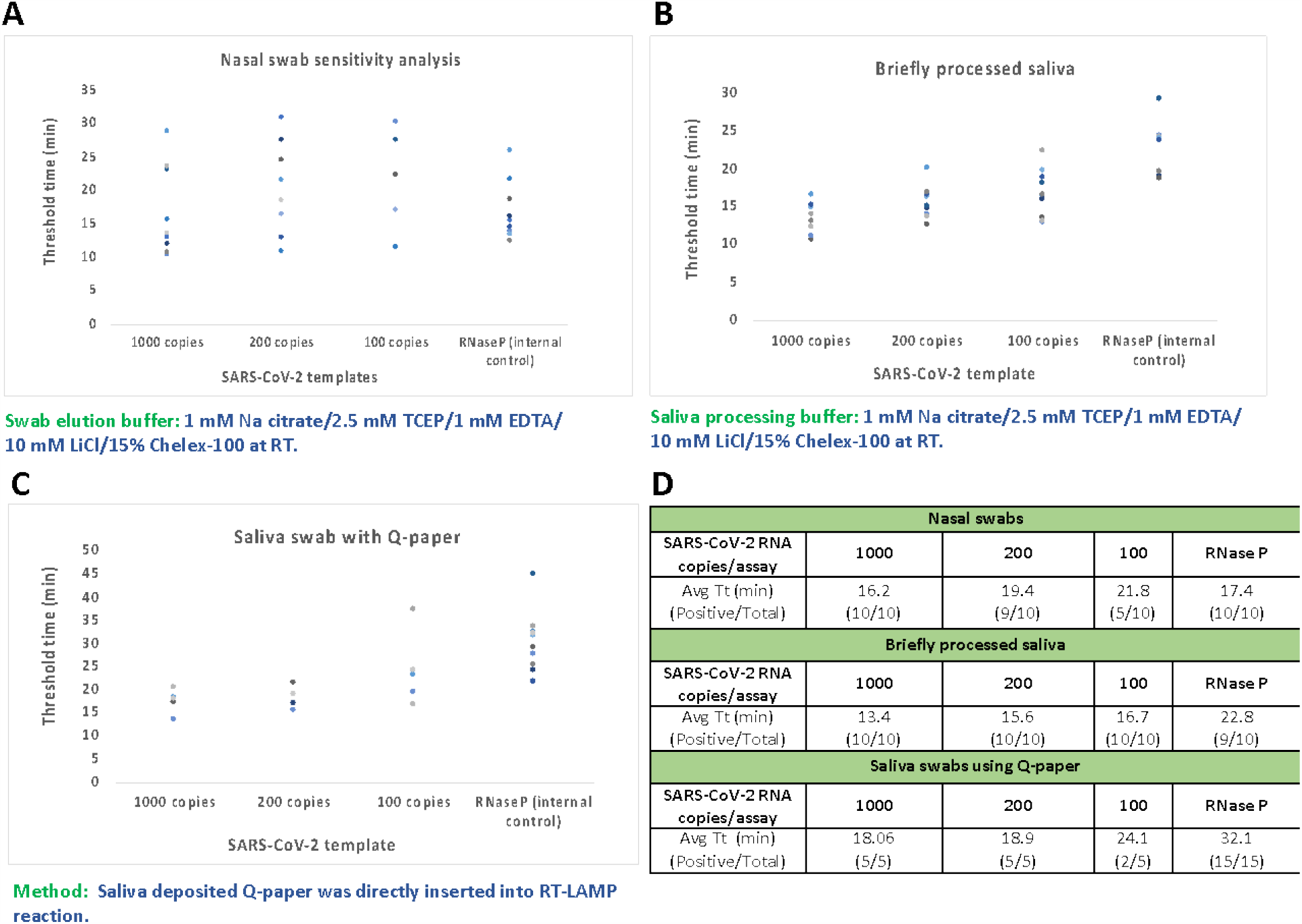
Further evaluation of presently preferred sampling methods and sensitivity analysis with contrived samples using heat-inactivated SARS-CoV-2 template from BEI. **(A)** Varying amounts of RNA was spiked into nasal swab samples from healthy individuals. 200 copies of RNA were detected with consistency and RNase P gene was used as sampling control.**(B)** and **(C)** Varying amounts of RNA was spiked into saliva samples or saliva that was deposited onto Q-paper, respectively. 200 copies of RNA were detected with consistency and RNase P gene was detected successfully. **(D)** Mean Tt values in minutes and numbers of positive results versus total number of samples were displayed in the table.

### Validation of DP-RT-LAMP assay with contrived saliva samples

Crude saliva was first added to RT-LAMP without any treatment, with a saliva: LAMP reaction mixture ratio of 1:5. As shown in **Fig 4B**, 1000 copies of RNA were detectable only after 40 min. This suggested that crude saliva was not very suitable as a sample on its own.

Suspecting that RNA might be rapidly degraded in saliva, saliva samples spiked with target DNA were tested (**S4 Fig**). Here again, the emergence of the signal was substantially delayed, even though the delay was not as large as with the analogous RNA. We then tried nasal elution buffers in more concentrated form. Here, saliva (100 µL) was treated with 100X buffer (0.25 M TCEP, 0.1 M EDTA, 0.1M NaOH or Na citrate, 1 µL) with or without 15% Chelex-100, and with or without a heating step. Multiple runs showed the most reliable results, with no signal in the absence of a template, with inactivation buffer containing TCEP, EDTA, sodium citrate, LiCl and Chelex-100 without a heating step; the LOD was ∼ 100 copies/assay. Fluorescent signals obtained from positive samples were clearly differentiable from those arising from samples lacking target (**Fig 4C**).

As an alternative to this saliva sampling method, saliva was absorbed on to Q-paper, which was placed after a brief time (5 min) at room temperature (to simulate how processing might occur in a workplace lobby) and directly added into the RT-LAMP mixture. Analogous to what is seen with mosquito carcasses[16], 100 copies of viral RNA could be detected by simply spotting SARS-CoV-2 RNA onto saliva coated Q-paper from which it was directly amplified by RT-LAMP. Visualization of the positive signals was obtained similarly using blue LED and orange filter combination (**Fig 4D**).

The sensitivities of the assay with two saliva sampling methods were further analyzed using contrived saliva samples of healthy volunteers. Here, 200 copies of RNA/assay could be detected by both methods with 100% detection rate with a small sample size (5 to 10 cases); 100 copies of RNA were detected with 100% efficiency using 100X inactivation solution, and with 40% efficiency using Q-paper for sampling. Additionally, the internal control targeting RNase P gene was detected at 90-100% in both methods (**Fig 5B, 5C and 5D**).

### Validation of DP-RT-LAMP assay on clinical samples

Previously collected nasopharyngeal (NP) swabs samples that were stored in viral transport medium (VTM) were first eluted at room temperature without a heating step in sample elution buffer containing sodium citrate, LiCl, TCEP, EDTA and Chelex-100. An aliquot of this elution was used for assay validation. SARS-CoV-2 specific primer set and internal control targeting RNase P gene was run in parallel and signal threshold times were determined. LAMP assay results were also compared to a CLIA notified and ICMR (Indian Council for Medical Research) approved PCR assay from GenePath Diagnostics (GPDx CoViDx One PCR). This is a 4-plex real-time RT-qPCR assay targets RdRP, N and E gene of SARS-CoV-2 and uses RNase P gene as sample extraction control. It uses a commercial sample extraction kit to purify and concentrate viral RNA from VTM whereas LAMP method uses minimal sample preparation through simple swab elution. Out of 11 samples tested, LAMP assay results are in 100% agreement with PCR assay with average Tt value of 17 min for SARS-CoV-2 target and 18 min for RNase P targets (**Table 2)**.

**Table 2.** Clinical evaluation of DP-RT-LAMP assay and comparable PCR assay.

| Sample | LAMP Tt (min) |  |  | GPDx CoViDx One PCR Ct |  |  |  |  |
| --- | --- | --- | --- | --- | --- | --- | --- | --- |
|  | CoV2-W3 | RNase P | Result | RdRP | N | R | RNase P | Result |
| 1 | 32.7 | 24.0 | Positive | 33.13 | 35.78 | 33.26 | 29.15 | Low Positive |
| 2 | 10.9 | 18.0 | Positive | 25.56 | 27.45 | 25.25 | 28.21 | Positive |
| 3 | 41.5 | 16.2 | Positive | 30.99 | 34.24 | 31.65 | 25.55 | Positive |
| 4 | 12.4 | 16.5 | Positive | 30.85 | 32.75 | 31.31 | 29.81 | Positive |
| 5 | 11.0 | 16.9 | Positive | 29.62 | 31.84 | 29.72 | 29.82 | Positive |
| 6 | 16.7 | 17.7 | Positive | 31.49 | 33.73 | 32.18 | 31.14 | Positive |
| 7 | 12.7 | 14.7 | Positive | 30.6 | 32.12 | 30.99 | 29.25 | Positive |
| 8 | 23.7 | 15.0 | Positive | 32.65 | 35.73 | 33.31 | 29.5 | Low Positive |
| 9 | 10.3 | 15.6 | Positive | 25.67 | 27.71 | 25.75 | 28.99 | Positive |
| 10 | 13.0 | 16.7 | Positive | 31.94 | 34.87 | 32.9 | 27.35 | Positive |
| 11 | 9.0 | 29.0 | Positive | 23.77 | 25.29 | 23.62 | 27.51 | Positive |

LAMP assay: Swab elution in 200 μL buffer. 5 μL input in 25 μL reaction
PCR assay: With RNA extraction (200 μL VTM input, elution volume of 35 μL), 5 μL Input 15 μL reaction.

### Multiplex detection of SARS-CoV-2 and RNase P

An assay robust for workplace use must incorporate a signal to indicate that sampling is sufficiently aggressive. Our displaceable probe architecture allows the simultaneous detection of viral RNA and the human RNase P gene in single tube. To show this, we spiked varying amounts of viral RNA into 440 copies of purified human RNA background. Using the LightCycler^®^ 480 and three fluorescent channels, 10 copies of SARS-CoV-2 RNA could be detected in the presence human RNA in two-plex format when equal amount of the two (virus and human) 10X LAMP primer sets were present.

When viral RNA was present in higher amounts, the signal for the positive control was delayed to 32.5 minutes, instead of appearing between 21-23 min. This is presumed to reflect the two amplification processes competing for some of the LAMP amplification resources. A similar degree of sensitivity for both targets was achieved when viral RNA was ∼ 1000 copies/assay (**Fig 6A**).

**Fig 6.**
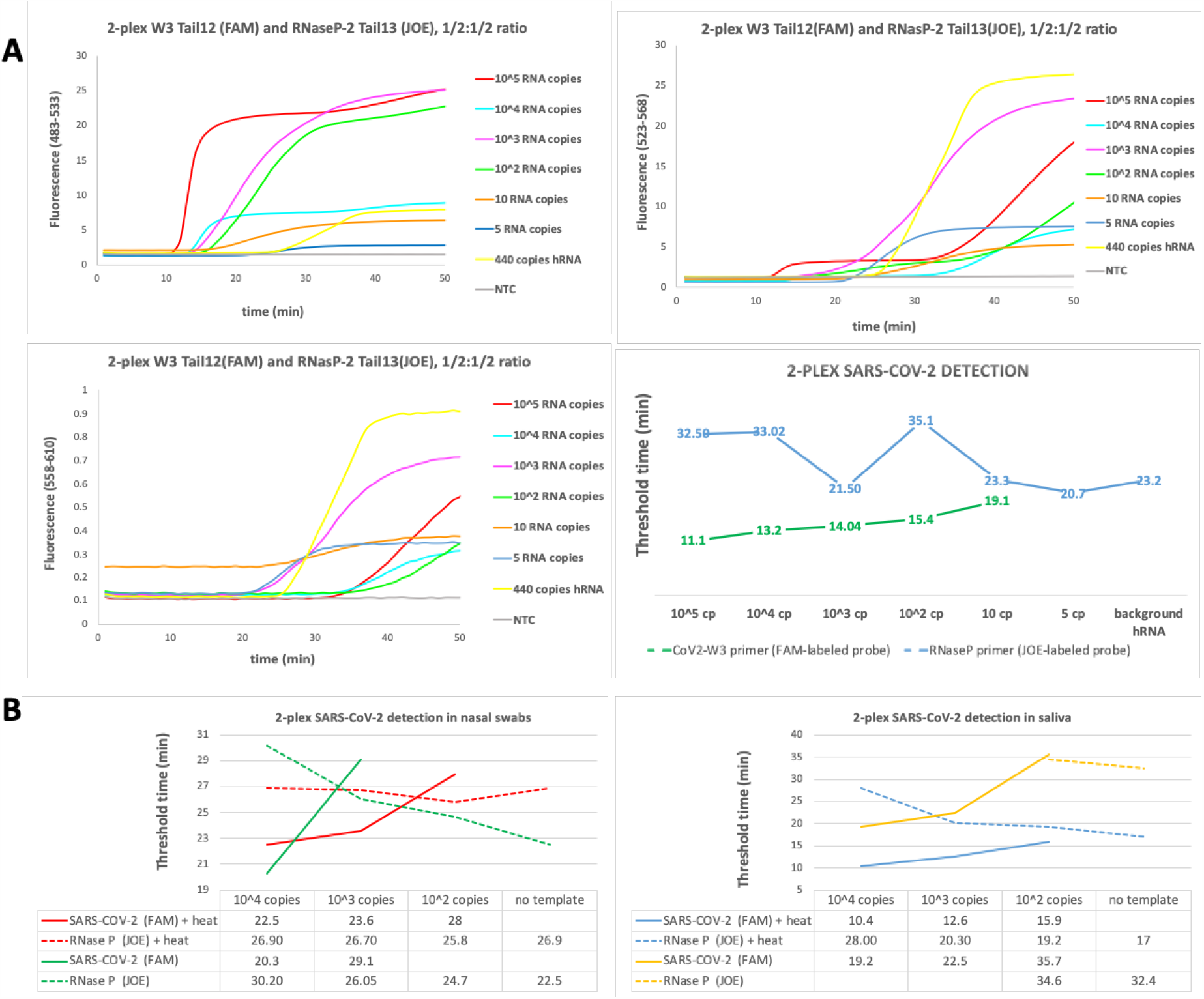
Multiplexed detection of SARS-COV-2 RNA and RNase P (internal control). **(A)** Varying amounts (10^5^, 10^4^, 10^3^, 10^2^, 10 and 5 copies) of heat-inactivated SARS-CoV-2 (BEI resources) spiked with 440 copies of purified human RNA. Fluorescence signals from three channels were recorded every 30 seconds using LightCycler^®^ 480. Channel 483-533 is specific for SARS-CoV-2 RNA, channel 523-568 can detect signals from both targets (ladder formation manifests itself), and channel 558-610 is specific for RNase P. Corresponding Tt values were shown on the table. **(B)** 10^4^, 10^3^ and 10^2^ copies of SARS-CoV-2 RNA was spiked into processed nasal swab and saliva samples and analyzed simultaneously. Effect of heating step was explored in 2-plex experiments and corresponding Tt values were shown on the chart.

We then spiked 10^4^, 10^3^ and 10^2^ copies of viral RNA into processed nasal swab samples and ran multiplexed assays. We also explored the effect of brief heating step at 95°C for 5 min on the assay sensitivity. When no heating step was involved, only 10^4^ and 10^3^ copies of viral RNA could be detected and Tt value of RNase P decreases with the decreasing order of viral RNA input. When samples were heated, more uniform co-amplification of both targets was observed with Tt ranging from 22 to 28 min for CoV-2 and 25 to 26 min for RNase P.

Similarly, we ran similar set of experiments for saliva samples using a brief processing step that is similar to nasal swab processing. When there was no heating, 10^4^ and 10^3^ copies of RNA could be detected within 19 and 22 min, but signal was substantially delayed to 35 min for 100 copies of RNA. When 10^4^ and 10^3^ copies of viral RNA were present, no RNase P signal was observed. This is expected of LAMP since the viral amplifications consumed LAMP resources in general (e.g. dNTPs). However, RNase P signal appeared at 35 min mark in the presence of 100 copies of RNA and at 32 min mark in the absence of viral RNA. When saliva samples were heated, more uniform co-amplification of both targets was observed with Tt ranging from 10 to 16 min for CoV-2 and 17 to 28 min for RNase P (**Fig 6B**).

### Lyophilization of DP-RT-LAMP reagents

For workplace entry use, the reagents used must robustly survive transport and storage in amateur hands. Accordingly, we prepared lyophilized reagent mixtures and tested their performance.

Here, the production process began by removing glycerol from the commercial enzymes as delivered. For this, an ultrafiltration column with a 10 kDa cut-off limit was loaded with DP-RT-LAMP enzymes and enzyme storage buffer was exchanged with the glycerol-free version. Then, 10X primers mix and dNTPs were added. The mixture was lyophilized for 4 to 6 hours, leaving dry reagents as a white fluffy powder (**Fig 7A**). Dry reagents were activated by rehydration buffer containing necessary salts and detergents and varying amount of viral RNA (1000 to 100 copies/assay). Alternatively, rehydration was done with contrived nasal and saliva samples (10,000 and 1000 copies of spiked RNA). Tt values were similar to their non-lyophilized versions with a few minutes of delay (**Fig 7B**) and clear visual fluorescent signals were recorded in all cases (**Fig 7C**).

**Fig 7.**
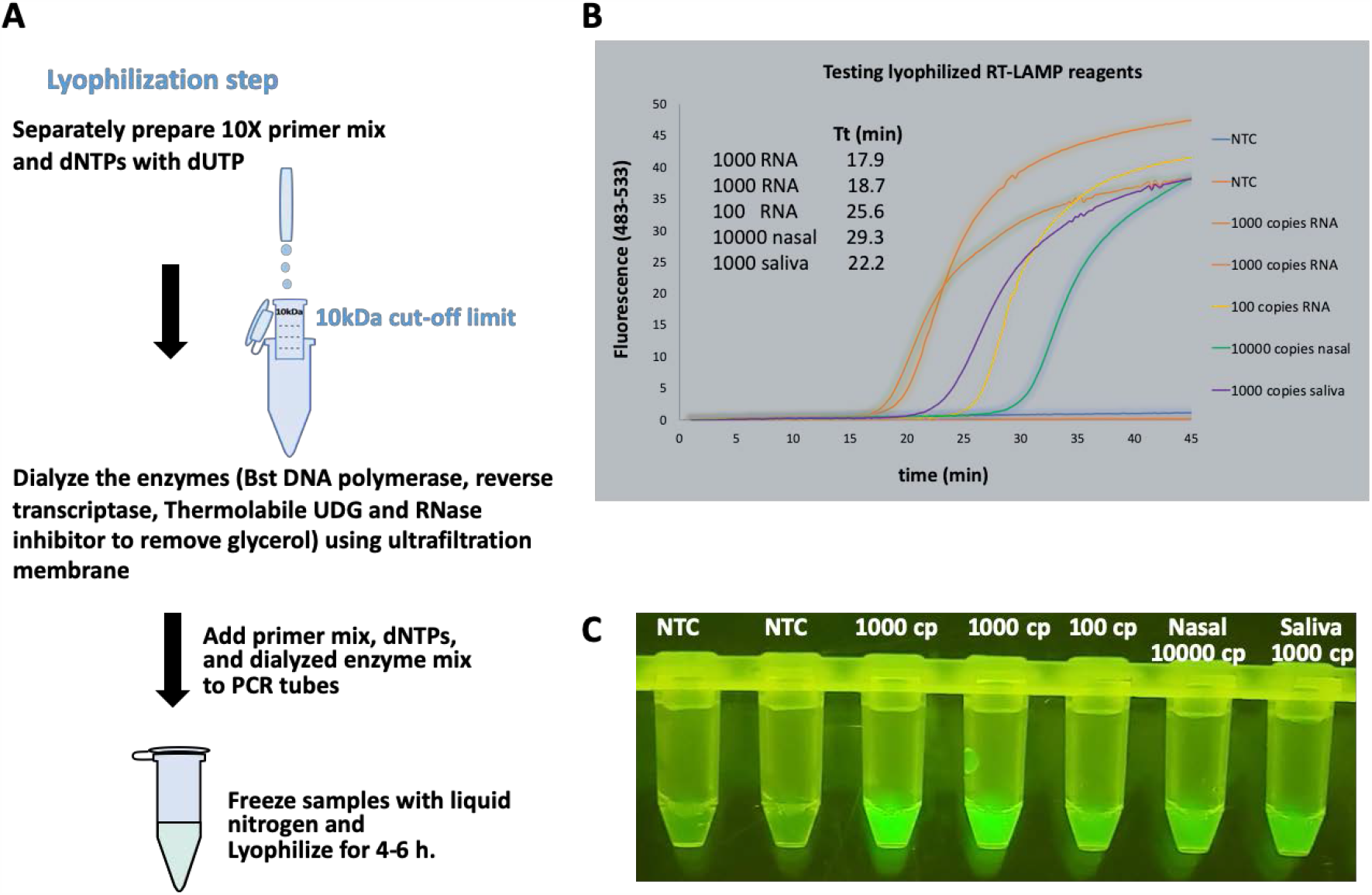
Lyophilization work-flow and analysis of dry reagents. **(A)** Workflow of lyophilization first involves the removal of glycerol from commercial enzymes. This was done by replacing enzyme storage buffer with its glycerol free version via ultrafiltration. The next step combined 10X primer mix and dNTPs with dialyzed enzymes. The mixture was then frozen (liquid N2) and lyophilized for 4-6 hours. **(B)** Lyophilized reagents were activated by supplementing lyophilized reagents with rehydration buffer, and templates containing SARS-CoV-2 RNA or contrived nasal/saliva samples; the DP-RT-LAMP progress was analyzed on Genie II and Tt values were determined. **(C)** End-point fluorescence was visualized using blue LED and orange filter.

## Discussion

The SARS-CoV-2 has been especially successful in having itself transmitted worldwide by its ability to cause a wide range of symptoms, ranging from lethality to no recognizable symptoms at all. Further, multiple anecdotal examples show clear transfer of the virus from one symptomatic patient to another symptomatic patient via an individual who displayed no symptoms at all, neither at the time of transmission nor forever after [48]. This makes SARS-CoV-2 virtually unique among pathogens, while presenting an unprecedented challenge to those charged to manage the resulting pandemic. While classical 2003 SARS could be tracked by monitoring symptoms (e.g. fever, which can be remotely tested), SARS-CoV-2 cannot.

This makes the current testing regime totally inadequate. Reports from hospitals and New York City, for example, show the time between sampling, sample transport, and return of sample results ranging from two days to 22 days, with costs per assay generally in excess of $100. Neither this time of delay nor the cost are adequate to determine whether or not the individual being sampled is likely to forward infect other individuals at a workplace, courtroom, jail, or schoolhouse. Indeed, as Thomas Frieden of the CDC pointed out, if the test requires days to resolve an outcome, the test might as well not be taken at all, except for curiosity or for larger epidemiological public health [49].

Compounding this is the fact that the asymptomatic carrier has no particular reason to present himself/herself for testing. The most common way in which such asymptomatic carriers are identified is by broad testing of individuals who have been in contact with asymptomatic patient. However, for serious epidemiological work to be done given the peculiarities of this particular virus, large-scale random testing must be done. Again, the standard procedures involving days to result and $100 per assay are not compatible with this.

It is easy to identify preferred workplace assay; however, it must cost just a few dollars to run. Further, the assay must be run without the need to transport samples. Essentially all “home assay” kits are not home assays at all, but rather are home sampling kits.

Further, the results must be returned in minutes, not hours, and certainly not days. Rather, the assay must be usable at the entrance to a workplace, a courtroom, an airport, or schoolyard, and return results in 30 minutes. If the assay is positive, the individual is referred off campus to a reference laboratory. If the assay is negative, to the extent that it does not indicate a risk for forward infection, the individual is allowed to enter the public space.

These specs make certain demands on assay design. First, they must not involve any of the classical sample preparation tests that are used in assays, and reference laboratories. They must be workable by a nonprofessional who need not be licensed, in an environment that must not need CLIA certification. Indeed, they cannot involve a reference laboratory at all.

These demanding specifications may be offset in part by the absence of a need for ultra-high sensitivity. For example, when treating HIV, even 10 virions in a sample indicate that the patient has an infection that demands medical attention. For workplace use, however, an assay need be only as sensitive as necessary to ensure that the individual does not present a forward contamination risk. While the viral load of saliva necessary for that risk is not known, emerging data suggest that this requires hundreds of thousands or millions of viral particles per milliliter for upper respiratory track samples [50].

The assay presented here meets all the requirements for use it in an entrance to a public space, such as a schoolyard, a workplace, or an airport. It detects virus if it is present at approximately 200 copies per nasal swab assay, representing approximately 8,000 copies of RNA per nasal swab, and 100 copies per saliva assay, representing approximately 20,000 copies of RNA per mL of saliva. This is currently believed to be below the level of mean viral load in upper respiratory specimens [51, 52] and below the level required for a forward infection risk [50, 53].

## Data Availability

All data presented in the manuscript and supplemental information file is available upon request.

## Acknowledgements

The following reagent was deposited by the Centers for Disease Control and Prevention and obtained through BEI Resources, NIAID, NIH: SARS-Related Coronavirus 2, Isolate USA-WA1/2020, Heat Inactivated, NR-52286.

## Author contributions

Conceptualization: Ozlem Yaren, Steven A. Benner Investigation: Ozlem Yaren, Steven A. Benner.

Methodology: Ozlem Yaren, Jacquelyn McCarter, Nikhil Phadke. Project administration: Ozlem Yaren.

Resources: Steven A. Benner. Software: Kevin M. Bradley.

Supervision: Ozlem Yaren and Nikhil Phadke.

Validation: Benjamin Overton, Zunyi Yang, Shatakshi Ranade, Kunal Patil, Rishikesh Bangale. Visualization: Ozlem Yaren, Jacquelyn McCarter.

Writing-Original Draft Preparation: Ozlem Yaren.

Writing-Review and Editing: Ozlem Yaren, Kevin M. Bradley, Nikhil Phadke, Steven A. Benner.

